# Releasing the lockdown in the UK Covid-19 epidemic: a stochastic model

**DOI:** 10.1101/2020.04.28.20083329

**Authors:** Anthony Lander

## Abstract

**Background:** In a classic epidemic, the infected population has an early exponential phase, before slowing and fading to its peak. Mitigating interventions may change the exponent during the rising phase and a plateau can replace a peak. With interventions comes the risk that relaxation causes a second-wave. In the UK Covid-19 epidemic, infections cannot be counted, but their influence is seen in the curve of the mortality data. This work simulated social distancing and the lockdown in the UK Covid-19 epidemic to explore strategies for relaxation.

**Methods:** Cumulative mortality data was transposed 20 days earlier to identify three doubling periods separated by the 17^th^ March—social distancing, and 23^rd^ March—lockdown. A set of stochastic processes simulated viral transmission between interacting individuals using Covid-19 incubation and illness durations. Social distancing and restrictions on interactions were imposed and later relaxed.

**Principal Findings:** Daily mortality data, consistent with that seen in the UK Covid-19 epidemic to 24^th^ April 2020 was simulated. This output predicts that under a lockdown maintained till early July 2020, UK deaths will exceed 31,000, but leave a large susceptible population and a requirement for vaccination or quarantine. An earlier staged relaxation carries a risk of a second-wave. The model allows exploration of strategies for lifting the lockdown.

**Interpretation:** Social distancing and the lockdown have had an impressive impact on the UK Covid-19 epidemic and saved lives, caution is now needed in planning its relaxation.

**Funding:** Unfunded research.

**Research in context:** *Evidence before this study:* The classical Susceptible, Infected, Recovered, (SIR) epidemiological model with additional compartments and sophistications have been widely used to make forecasts in the Covid-19 pandemic but are not easily accessible.

*Added value of this study:* This study adds reassurance that the interventions of social distancing introduced on the 17^th^ March and the lockdown of the 23^rd^ March 2020 have reduced mortality. The risks of a second-wave on their relaxation are real and illustrated graphically.

*Implications of all the available evidence:* Together with other models, credence is given to the risks of a second-wave in the UK Covid-19 epidemic on the relaxation of restrictions.

## Introduction

In response to the Covid-19 pandemic, nations have imposed social distancing of 2 metres, and reduced the number of social interactions in lockdowns with significant economic and social consequences. The restrictions ‘flatten the curve’ of infections. Choosing how to lift restrictions is not easy. An early relaxation could initiate a second-wave.

The doubling period in mortality in the global pandemic was initially around 6 days but more recently in Europe it has been higher at around 4 days, see Figure 1.

**Figure 1.**
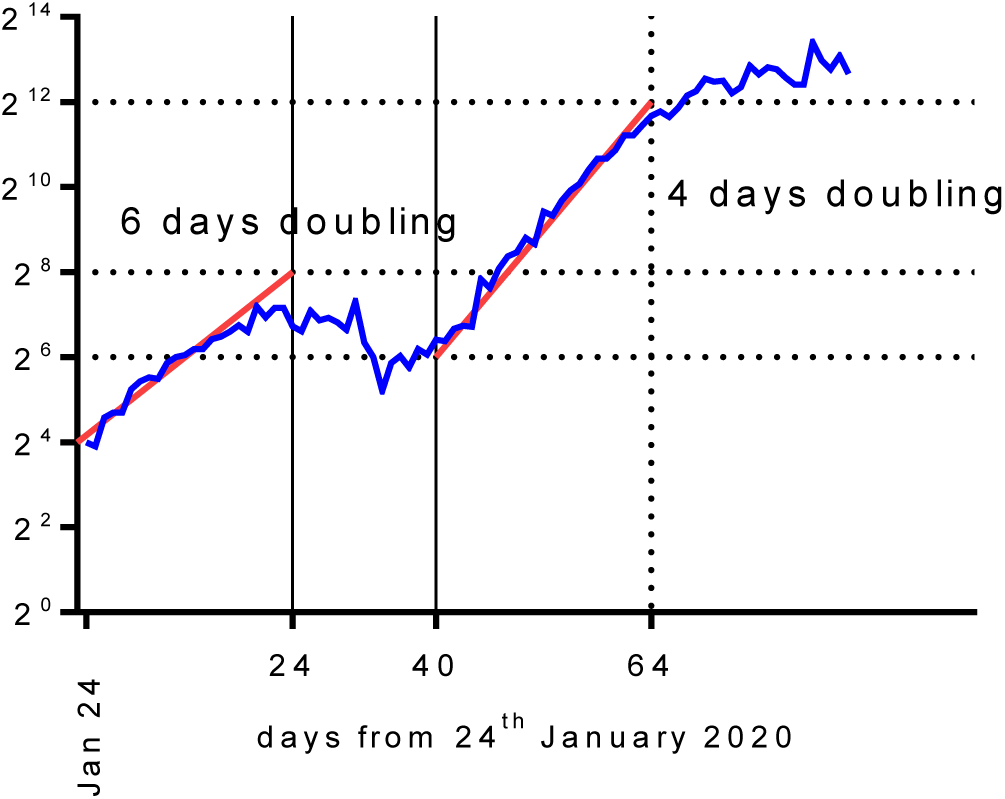
Global deaths from Covid-19 with a logarithmic axis showing doubling times (worldometers.info).

On the 17^th^ March with a forecast of 260,000 UK deaths social distancing was introduced (1) followed by the lockdown which started on 23^rd^ March. UK Covid-19 deaths from 15^th^ March were initially doubling every 3 days, Figure 2.

**Figure 2.**
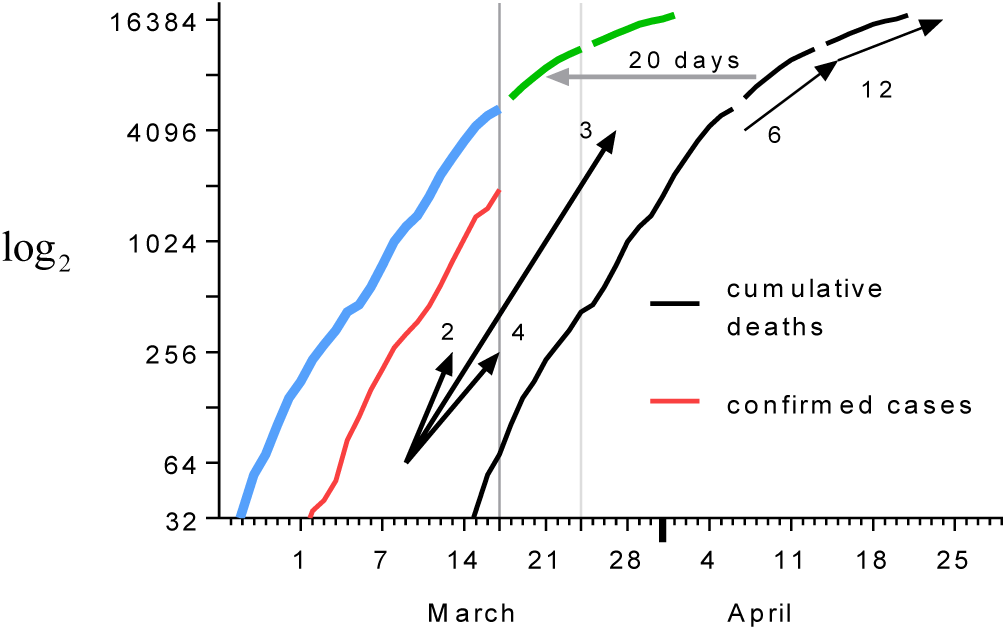
UK cumulative deaths (black) from 15^th^ March and transposed 20 days earlier (blue, green). The *log*_2_ scale makes doubling easy to see, arrows show gradients for doubling every 2, 3, 4, 6 and 12 days. Data from worldometers.info.

In Figure 2, when the curve for cumulative deaths is brought forward 14 days it is similar to the curve for confirmed cases. This tells us that the deaths arose 14 days after infections were confirmed but not about the interval from acquiring infection. The curve for cumulative deaths has been transposed 20 days earlier and has a relationship with the introduction of social distancing 17^th^ March and the lockdown starting on the 23–24^th^ March. The doubling time in mortality then rose from 3 days to 6 days, then 12 days.

The following are unknown; (a) the proportion of the population currently infected on any day and (b) the proportion of the population acquiring an infection on any day. These are related, and before immunity or death, the proportion currently infected is simply the sum of the proportions who acquired an infection each day up to that point in time. Up until that point in time, if the number of people acquiring an infection each day doubles every three days, then the number of people currently infected will likewise also double every three days: the integral of the exponential function being an exponential function with the same exponent.

If a fraction of those who are infected die after some fixed interval, then after that interval, daily deaths will also double every three days when deaths first occur. As a first approximation the exponent of the exponential phase of the rise in daily deaths, which we know, tells us about the exponent of the exponential phase in both the proportion of the population acquiring infection and in the proportion of the population infected on a daily basis at the start of the epidemic—neither of which we know. When we consider the variance in that interval to death, we see that the link between these exponential curves will be less faithful. However, the variance will increase the doubling time (decrease the exponent) in deaths in comparison with the doubling time in the acquisition of infection. This is helpful to us.

It is self-evident that if the peak proportion of the infected population is high then the risk of a second-wave is low, and if the peak proportion is low there is a higher risk of a second-wave. For this reason, a margin of safety is added if we use the known exponent of the exponential phase in mortality for the unknown exponent for the exponential phase in the curves representing infections.

Preliminary WHO data suggests that the median time from onset to recovery for mild cases is approximately 2 weeks, and 3–6 weeks for patients with severe or critical disease. Amongst patients who died, the time from symptom onset to death ranged from 2–8 weeks (2).

There is often great benefit in making explicit the assumptions of ones understanding of a real-world phenomenon, which changes in time, in a model which can be formulated mathematically in simultaneous partial differential equations, and then studying the solutions. For this reason, epidemics are often followed with a set of differential equations representing the size of the susceptible *S*, infected *I*, and removed or recovered *R* (dead + immune) populations. Modifications using related and more sophisticated compartments are also described and have been recently reported for Covid-19 (3).

In its simplest form, using positive constants *β* and *γ*, one imagines 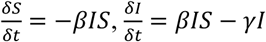 and 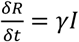. From these, the early infected population grows exponentially then flattens off, peaks and then falls whilst *S* falls and *R* rises. Though these equations, and their cousins, describe some of the features of epidemics they are deterministic whilst nearly all non-trivial real-world problems have outputs that vary because of stochastic processes acting during their evolution. Furthermore, real-world systems typically generate divergent outputs that are sometimes very sensitive to the initial conditions.

An attractive alternative to the descriptive partial differential equations is to represent each individual in the memory of a computer and then manipulate them according to explicit and modifiable rules whilst incorporating a number of stochastic processes. Though superficially simple, with seemingly naïve axioms and assumptions, stochastic models can demonstrate robustness and generate useful and stable outputs.

All models are wrong but some are helpful (4). But worryingly some could be unhelpful, especially if they wrongly inform public policy. When considering the Covid-19 pandemic we do not, at present, know how many people are currently infected or are asymptomatic but contagious. However, by transposing the mortality curves back 20 days we may assume that the early rise in infections was doubling every 3 days and that this changed on introducing social distancing and the lockdown.

Before building the stochastic model, the following features and expectations were considered to be manifestations of robustness and against which the model was tested and developed: —

1. Epidemics should have some stability in demonstrating similarly shaped curves in the number of infections over a majority but not all simulations for the same parameters. These outputs should, in the main and when averaged, be similar to SIR modelling.
2. The left-hand tail of the curve for infections should be the most variable feature over a number of simulations, especially if seeded with only one initial contagious individual.
3. Many stable curves should be found over a reasonably wide variation of input parameters, each allowing an infection to spread before dying out but without rapidly infecting all of the individuals. There should be epidemics that leave some individuals uninfected, alive and without immunity.
4. In contrast, there should also be some sets of parameters, under which there is sometimes an epidemic and sometimes no significant spread at all. These are the bifurcations one expects in chaotic systems.
5. There should be stable epidemics whose first peaks pass, leaving a sufficiently large susceptible population at risk of a second-wave.
6. Since this model was not going to include loss of immunity (which allows re-infection), birth or migration, endemic phenomena should not be expected.

## Methods

An easily modified stochastic model was written in Microsoft Excel VBA v7·1 operating in Windows 10. Rather than having an *R*_0_ dictate the early exponential growth in the infected population, or a set of differential equations to describe populations and compartments, these simulations used a number of stochastic processes representing biological and behavioural phenomena.

Individuals were assigned a number of characteristics including: age, sex, risk of mortality if infected, a measure of daily viral exposure, a susceptibility factor, an incubation period, and two contagious periods. One contagious period was for mild illness and the other for serious, critical or fatal illness. Each individual also had fields for location, symptom status, immune status, alive/dead, number of daily contacts, and two clocks; one for the individual’s assigned contagious period and one for a virtual ‘*twin*’ who was assigned a fatal illness contagious period.

Many viral illnesses have incubation periods which are lognormal, or something remarkably similar. A lognormal distribution was used based on Covid-19 data (5), see Figure 3.

**Figure 3.**
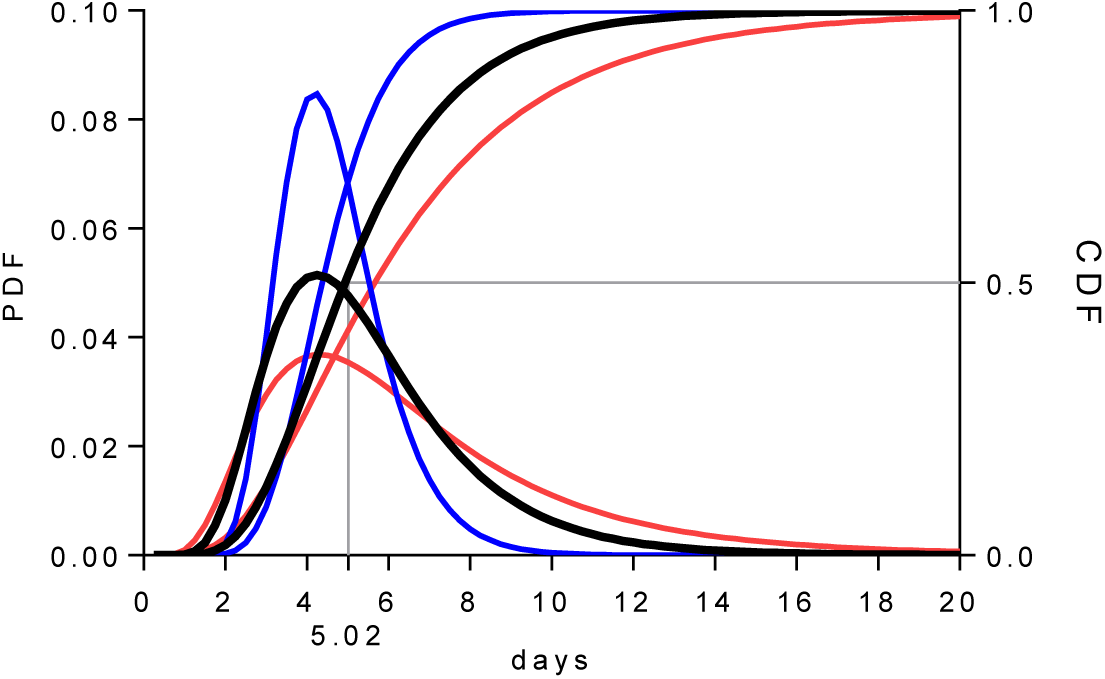
Lognormal distribution in black with its Probability Density Function PDF (left axis) and Cumulative Density Function CDF (right axis). In red and blue are the 95% confidence intervals for the distribution.

For these simulations the *meanlog* and *sdlog* were 1.621 and 0.418 respectively. Each individual was assigned a randomly chosen age and sex such that the population represented the demographics of the 2011 UK census.

Knowing the age and sex, an individual was designated to die if infected based on a probability taken from early Chinese data, released on February 11^th^ 2020, see Table 1.

**Table 1.** Preliminary estimates of the mortality rate by age and sex. February 11^th^ 2020 Chinese Centre for Disease Control and Prevention n=44,672.

| Age in years | Mortality | Male | Female |
| --- | --- | --- | --- |
| 10-19 | 0.002 | 0.0033 | 0.001212 |
| 20-29 | 0.002 | 0.0033 | 0.001212 |
| 30-39 | 0.002 | 0.0033 | 0.001212 |
| 40-49 | 0.004 | 0.0066 | 0.002424 |
| 50-59 | 0.013 | 0.02145 | 0.007879 |
| 60-69 | 0.036 | 0.0594 | 0.021818 |
| 70-79 | 0.08 | 0.132 | 0.048485 |
| 80+ | 0.148 | 0.2442 | 0.089697 |

There were two contagious periods which ended with immunity or death. Individuals being assigned a fatal outcome in the event of becoming infected were assigned a longer contagious period. A contagious period was assigned to most infected individuals as a lognormally distributed random variable with a *meanlog* of 2.639 and *sdlog* of 0.2. A random 5% were assigned to be critically ill, and together with the dying individuals, this group had contagious periods with a *meanlog* of 2.99 and *sdlog* of 0.223, see Figure 3.

**Figure 4.**
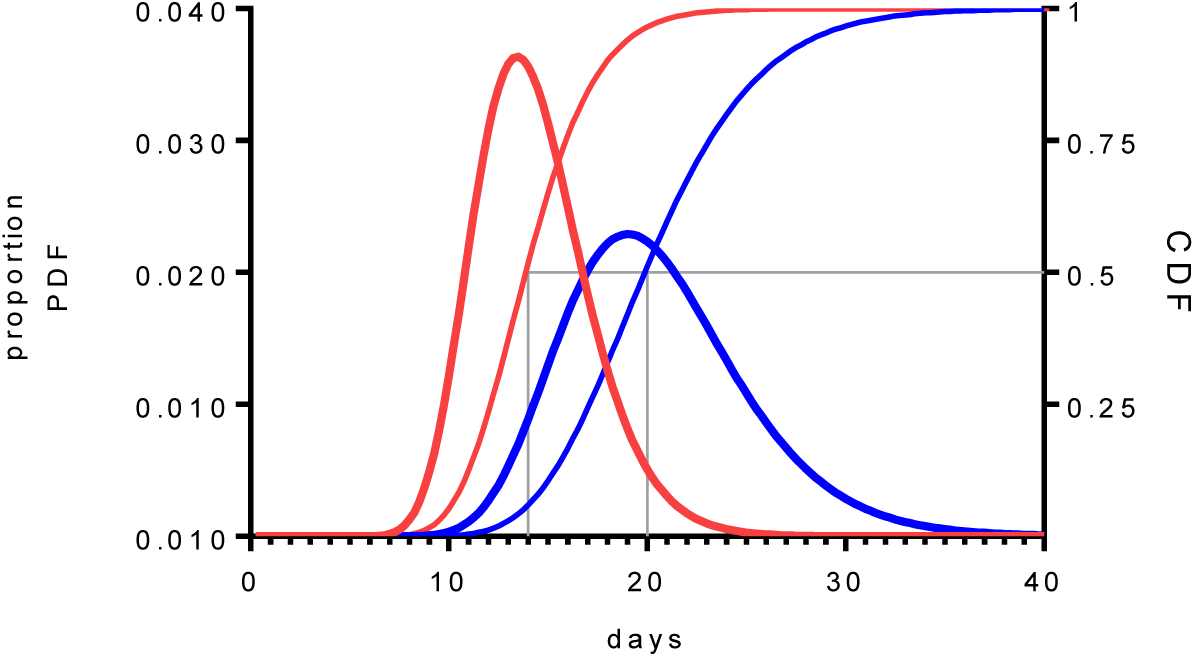
Probability density functions (left axis) and cumulative density functions (right axis) for the contagious period for most individuals in red, and the 5% critically ill and those who die in blue.

A susceptibility factor *s*, was randomly chosen such that it was normally distributed with a mean of 1 and standard deviation of 0.2. Randomly located and randomly moving individuals become infected if they exceed a daily viral load based on their cumulative proximity to contagious near neighbours and their susceptibility factor. The exposure, consequent upon an interaction between a susceptible individual *i* and a contagious individual *j*, had a relationship with the inverse square distance separating the individuals *d_i,j_*. All separations *d_i,j_* less than 0 2 metres were assumed to be at 0·2 metres. Separations exceeding 5 metres were ignored. When social distancing was applied, separations less than 2 metres were treated as if at 2 metres, this rule was applied for a percentage *x* of the interactions and is indicated as *SD_x_*. Thus *SD*_50_ means that 50% of interactions achieved social distancing and *SD*_10_ means that only 10% of interactions achieved social distancing. A threshold *t*, determined the daily cumulative viral load required for infection such that: for any susceptible *i* on any day, if 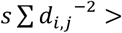 then *i* became infected. It was found that *t* = 1 permitted epidemics with appropriate population densities.

A period of one day was allowed to elapse between infection and symptoms. A percentage of symptomatic individuals made no movements, but still interacted with individuals that came within 5 metres. These individuals stayed at their local origin representing some measure of isolation. The percentage of symptomatic individuals confined is designated *C_%_*. Those not confined, though designated as symptomatic, represent asymptomatic viral spreaders since they are fully mobile.

Individuals moved *n* times a day and designated *M_n_*, as in a random walk, with step-size being 10 times the output of an inverse cumulative normal distribution Φ^−1^ (mean 0 metres, sd 10 metres). Individuals returned to their local origin at the end of each day. Mobile individuals were not allowed to leave the boundaries of the square, with some movements being reflected at the boundaries.

In these simulations 2000 individuals were each assigned a random local origin, in a square with sides of 490 metres giving an average population density of 8,330 km^−2^. In UK cities, residential population densities are in the region of 1,000–15,000 km^−2^ but social interactions in travel, work and recreation mean that far greater local densities are experienced for protracted periods. Only small populations were needed since each individual’s twin was being followed with a critically ill contagious period to death to simulate mortality data. The 2000 were sufficient to generate robust SIRI like epidemiological curves. One or more randomly placed contagious individuals were used to ignite an epidemic.

### Development

The principal outputs were the daily numbers of susceptible, infected, immune and dead individuals. Secondary outputs included metrics about interactions and checks on the incubation period distribution, demographics and mortality rates against age and sex (not reported here). In early simulations the mortality figures were a function of the demographic data and had a mean that followed a curve that mimicked, but was smaller than and lagged in time behind, the curve representing the number of infected cases. For this reason, the mortality output incorporated significant and uninformative noise.

Since the mortality from Covid-19 is probably less than 1%, and in the model, mortality had a low probabilistic link to the demographics, there was more information encoded in the infection output than the mortality output. In order to examine the fidelity of the link between the exponential phase in mortality and that in infections, and to overcome the noise inherent in the sparse numbers of modelled deaths, each individual was assigned a *‘twin’* who was given a critically ill contagious period, and after the individual had recovered and was immune, their *‘twin’* was followed to their death. The twin did not influence the curve of current infections or the behaviour of the epidemic. In this way the variance in the critically ill contagious period was allowed to influence the exponential phase in a larger virtual mortality data set. The virtual twins became the output of a virtual, and much larger epidemic (at least 100 x larger), whose infected population did not need to be simulated.

The stochastic nature of the model was examined. With lower density populations the first few contagious individuals could wander around for a variable number of days before igniting an epidemic. This phenomenon could delay the peak of infection for up to 10 weeks. This is an important observation. If there were a number of relatively isolated regional populations each of which suffered an epidemic peaking at a different time it does not follow that they were seeded at different times or that infection spread from one population to another in some defined sequence. Indeed, one region with a later inoculation may manifest its epidemic well before another region that was inoculated earlier.

Sometimes a set of parameters, that reliably initiated a set of similar epidemics, would occasionally fail to ignite an epidemic and instead infect only a small number of individuals. In contrast, no set of parameters was found that repeatedly generated epidemics which left a reasonably sized susceptible population whilst occasionally generating a runaway epidemic that infected all of the population in a very short period of time. This was reassuring real-world like behaviour.

With certain, easily found parameters, epidemics with approximate doubling of infected cases occurring every 3–4 days were obtained. Repeated simulations were performed for each set of parameters. This allowed an estimate of the variance in the output arising from the stochastic processes.

### The Covid-19 parameters

The Covid-19 epidemic simulations were seeded with 5 randomly placed contagious individuals to encourage similar, and short, left-hand tails for the simulated infected populations. With five seeds the exponential phases differed by no more than 3 days. This meant that the susceptible populations were similar sizes once lockdown was initiated.

Simulations started with 18 movements a day (*M*_18_) and social distancing was achieved for 10% of interactions (*SD*_10_), with 90% of the symptomatic individuals being confined to their local origin (*C*_90_). These parameters established an epidemic with an exponential phase having a doubling time of around 3 days. This phase ran until the susceptible population had fallen to 91%.

Social distancing restrictions were then imposed to increase the doubling time on the rising phase of the epidemic to mirror that seen in the UK mortality data shown in Figure 2. Social distancing was modelled such that 50% of interactions separated by less than 2 metres were treated as if at 2 metres (*SD*_50_) and movements were reduced from 18 to 14 a day (*M*_14_). The fraction of symptomatic individuals confined to their local origin was maintained at 90% (*C*_90_). These represented the real-life changes initiated around the 17^th^ March.

When the susceptible population fell below 79% the lockdown was modelled. The interval from the first mitigation on the 17^th^ March to the 79% susceptible level was within 6 or 7 days, bringing us to the lockdown starting on 23–24 March. Lockdown involved reducing movements from 14 to 7 a day (*M*_7_), increasing the achievement of social distancing to 60% of interactions *SD*_60_, and confining all symptomatic individuals to their local origins (*C*_100_).

The parameters for the epidemic were as follows: {*M*_18_, *SD*_10_*, C*_90_} to 17^th^ March, then the first mitigation was social distancing with a small reduction in movements {*M*_14_*, SD*_50_*, C*_90_} which ran to the lockdown starting on 23–24 March when the parameters became {*M*_7_*,SD*_60_, *C*_100_}.

Eight simulations of the UK Covid-19 epidemic appear in Figure 5 in red, blue and green for the infected population in its three phases (right axis). The eight grey curves are the modelled daily mortality (left axis). The actual UK daily mortality data is shown in the grey histogram topped with a black line.

**Figure 5.**
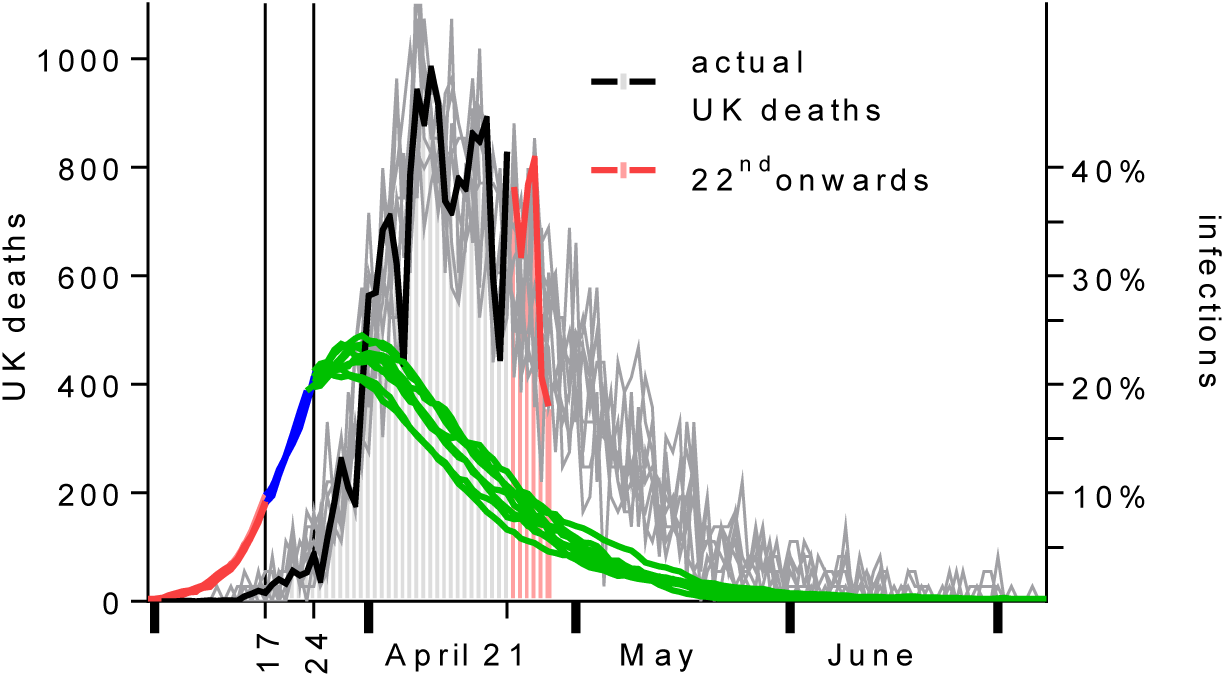
Eight simulated epidemics. Infected populations (right-axis) in red until social distancing (17^th^ March), in blue until lockdown (24^th^ March), and then in green. Simulated mortality, with marked daily variations is shown in grey (left-axis). Superimposed is the actual daily UK mortality (worldometers.info) plotted to the 21^st^ April in black, when the graph was first plotted. Additional data for deaths on 22–27 April have been added in red.

In Figure 5, daily mortality peaks on 9^th^ April and the epidemic ends in early July. The simulated daily mortality amongst the 2000 twins had the same shape as the actual daily mortality to 21^st^ April. If the simulated mortality was magnified by a factor of 27·5 the two were superimposable. Subsequent actual deaths to the 27^th^ April have been added in red and lie within the band of the simulated daily mortalities.

The cumulative UK mortality was 20,733 on the 26^th^ April 2020, and by that date 66% of the simulated deaths in the locked down epidemic had occurred. This suggests that for the UK, total deaths might reach 31,200.

In the locked down simulated epidemics roughly 1070 (54%) of the 2000 simulated individuals acquired an infection. If this proportion can represent 54% (34.8 million) of the UK population and if 31,200 of this group dies, then the case fatality rate is about 1/1000. This is a tenth of the estimate produced by the Chinese Centre for Disease Control and Prevention, see Table 1.

The simulations were averaged and plotted using a *log*_2_ axis, see Figure 6 for comparison with Figure 2. The susceptible population falls to just below 50% and is plotted on a linear axis on the right.

**Figure 6.**
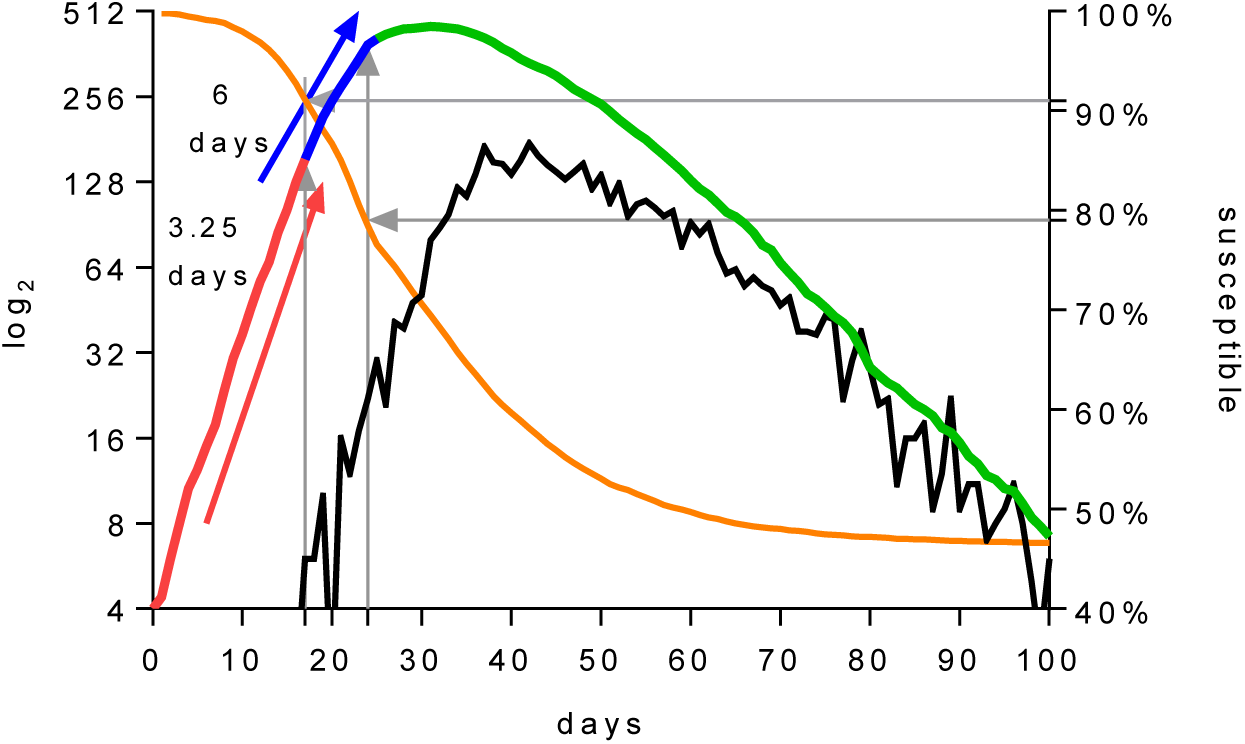
An average of the simulations, shown in Figure 5, are plotted against a *log*_2_ axis. The doubling time is 3·25 days (red) before the introduction of social distancing, and 6 days thereafter (blue) until the lockdown (green). The logarithm of the daily mortality of the simulations is shown in black. The susceptible population is plotted on the right axis.

### Examining the influence of the two distributions of the contagious periods

Five percent of infected individuals, and those who died, were given a different distribution for their contagious period, see Figure 4. The 5% figure was arbitrarily chosen and its impact on the behaviour of the simulations was examined. Simulations show that the impact on the overall curve of infections was small in comparison to stochastic variations, though there was an influence on the mortality curve, see Figure 7 left panel. The distribution of critically ill periods influences the shape and timing of the daily mortality curve.

**Figure 7.**
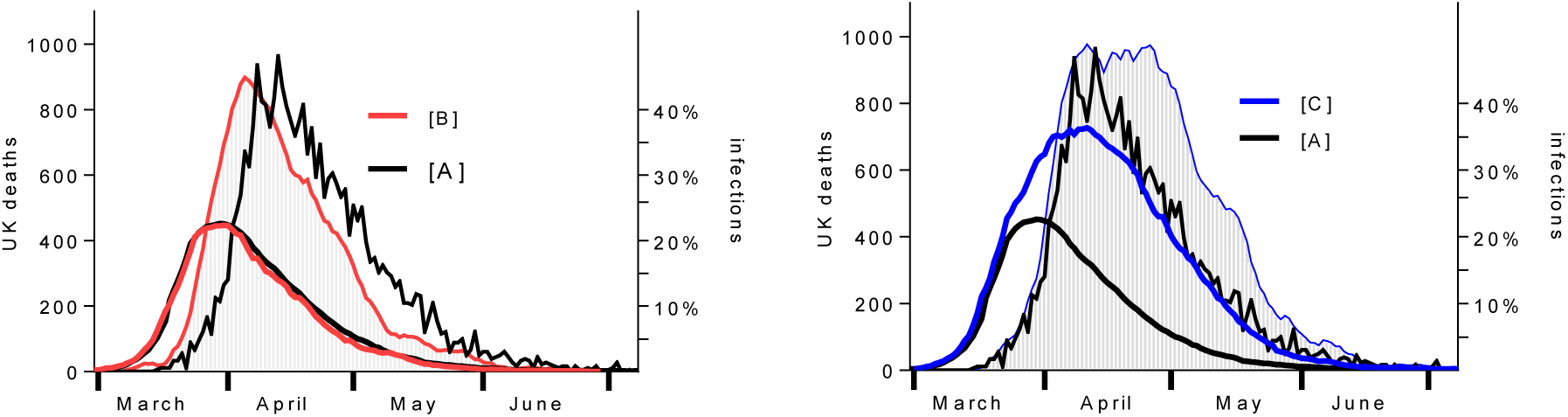
Two simulations of epidemics shown against the averaged simulations taken from Figure 5 in black. The black curves use two contagious periods, one for mild illness and one for the critically ill. [B], uses only the short contagious period (for mild cases) for all individuals, and [C] uses the longer contagious period (for the critically ill) for all individuals. The mortality curves for [B] and [C] have been smoothed for clarity

In Figure 7, simulations [B] and [C] compare the behaviour of using only one contagious distribution to influence the epidemic and its curve of infections. The periods are those shown in Figure 4. Normally the critically ill distribution applies to only 5% of individuals. As anticipated, in [A] the curve for infections is relatively insensitive to the 5% proportion having a longer contagious period whilst the mortality curve is shifted by about 1 week. In [B] all infected individuals had a contagious period from the critically ill distribution and the curve for infections was then markedly altered. It is the change in the curve of infections which leads to the mortality curve in the right-hand panel being larger and displaced to the right. These findings tell us that the critical ill fraction, and the distribution of their contagious periods is important in generating the shape of the daily mortality curve.

In the following simulations the epidemic always ran with the same parameters used for Figure 5 namely *{M*_18_*, SD*_10_*, C*_90_*} → {M*_14_*, SD*_50_*, C*_90_*} → [M*_7_*, SD*_60_, *C*_100_] with the transitions taking place on the 17^th^ and 23-24^th^ March. The infections are shown in black against the right axis and the daily mortality, being the average of a number of simulations, is shown in black against the left axis.

### Relaxing the lockdown

The first relaxation of a lockdown has movements alone increasing on the 20^th^ April, whilst maintaining social distancing, but at 50%, and maintaining confinement of the symptomatic. In Figure 8 the lockdown was released on 20^th^ April to assume the parameters *{M*_18_*,SD*_50_, *C*_100_}.

**Figure 8.**
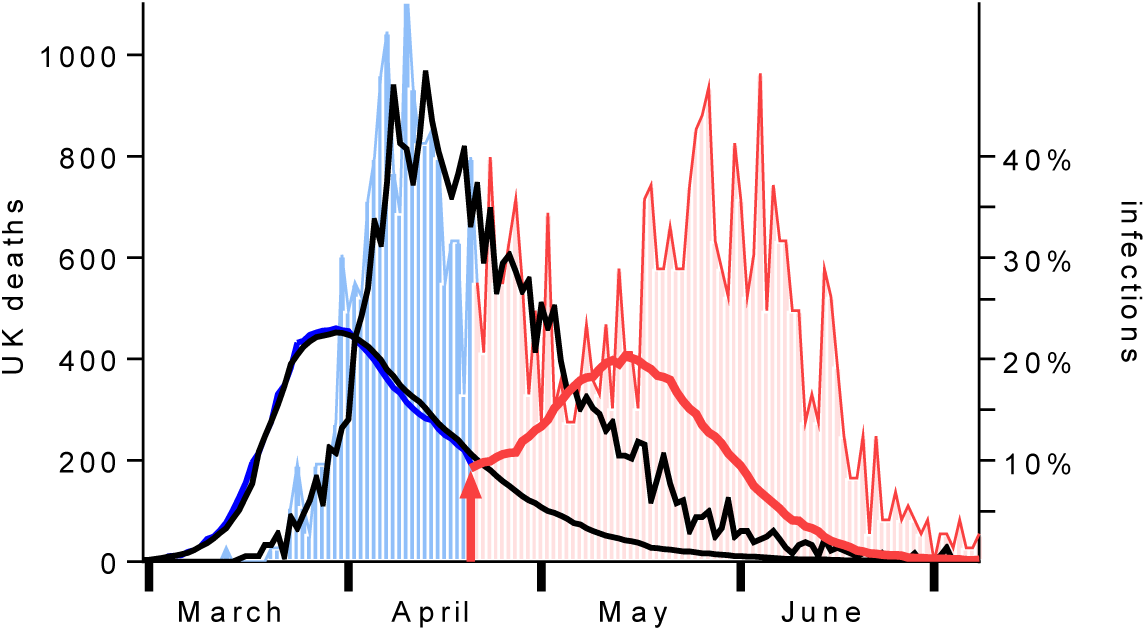
Averaged locked down simulations in black. A locked down epidemic is simulated (blue) and is consistent with previous simulations in black. The lockdown is released (red arrow) on the 20^th^ April, whilst maintaining social distancing. There is a second-wave in infections in May and a second-wave in mortality in late May and June (red). The second peaks are similar to the first peaks seen in March and April.

In Figure 8 second-waves are seen both in infections and later in mortality. This would be a problem for services. The ratio of simulated mortalities was 1821/1071 which represents 53,000 UK deaths, more than 21,000 additional premature deaths compared with maintaining the lockdown.

The next simulation is of a relaxation on the 1^st^ May and is shown in Figure 9. Movements were relaxed above the original 18 to 20, only 10% of interactions achieved social distancing and only 75% of symptomatic individuals were confined to their original location *{M*_20_*, SD*_10_*, C*_75_*}*. This represents a post-lockdown increase in social activity, which is a risk.

**Figure 9.**
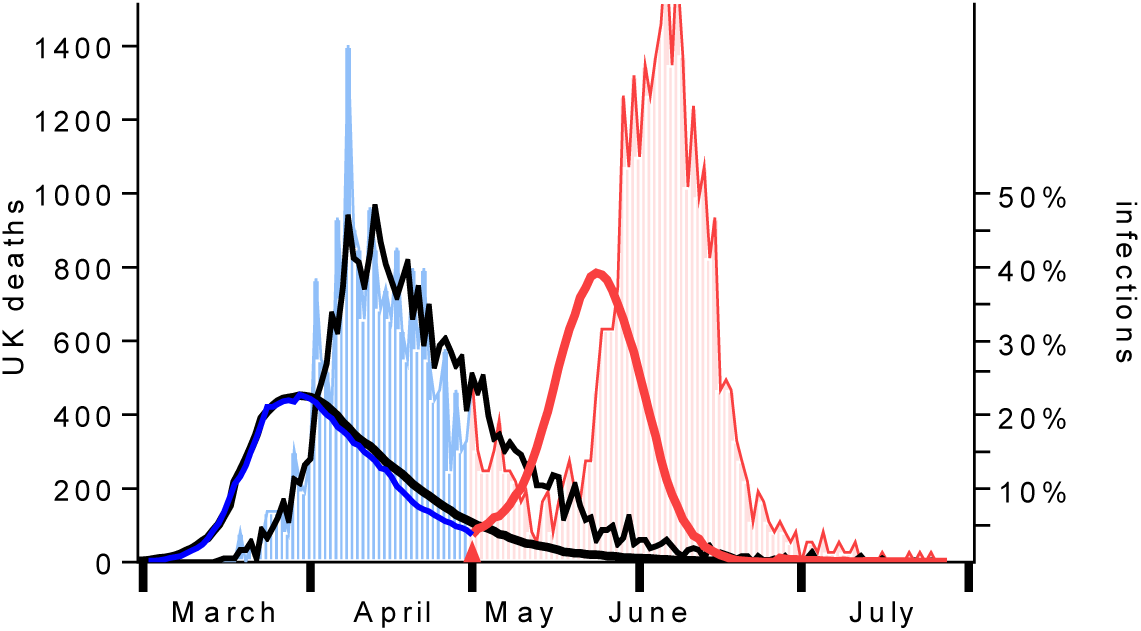
Averaged simulated locked down epidemics in black without relaxation. An additional epidemic is shown in blue, consistent with previous simulations until the release of the lockdown in red after the 1^st^ May. There is a peak in infections rising to 40% of the population and daily deaths reaching a peak of over 1400 in early June.

In Figure 9 the impressive second-wave would be a problem for services. There would be additional deaths. The ratio of simulated mortalities was 1985/1071 which represents 58,000 UK deaths.

In the simulation shown in Figure 10 the lockdown was relaxed later on the 10^th^ May with the same parameters {*M*_20_, *SD*_10_, *C*_75_}.

The mortality associated with Figure 10 is 56,500. If we compare these two similar strategies, whose lockdowns were relaxed at different times, there is less of an impact on relaxing restrictions on the 10^th^ May than on the 1^st^ May, but both would represent unacceptable outcomes.

**Figure 10.**
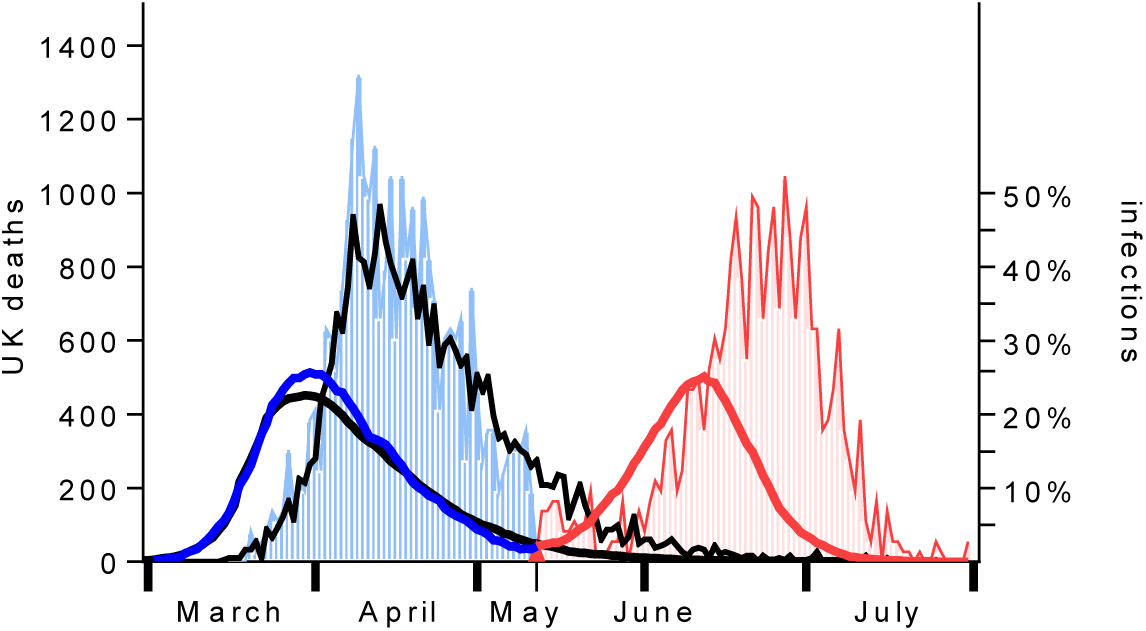
Averaged simulated locked down epidemics in black without relaxation. An additional epidemic is shown in blue, consistent with previous simulations until the release of lockdown in red after the 10^th^ May. There is a peak in infections rising to 25% of the population and daily deaths reaching a peak of over 800 in late June.

We next move to the 21^st^ May for two strategies which maintain social distancing whilst keeping the symptomatic confined. In one there are 20 movements and in the other 18 movements.

In Figure 11 there is relaxation of the lockdown on the 21^st^ May which maintains social distancing but allows movements to return to pre-epidemic levels *M*_18_ or exceed them slightly *M*_20_, the associated deaths equate to 48,600 and 50,100 respectively.

**Figure 11.**
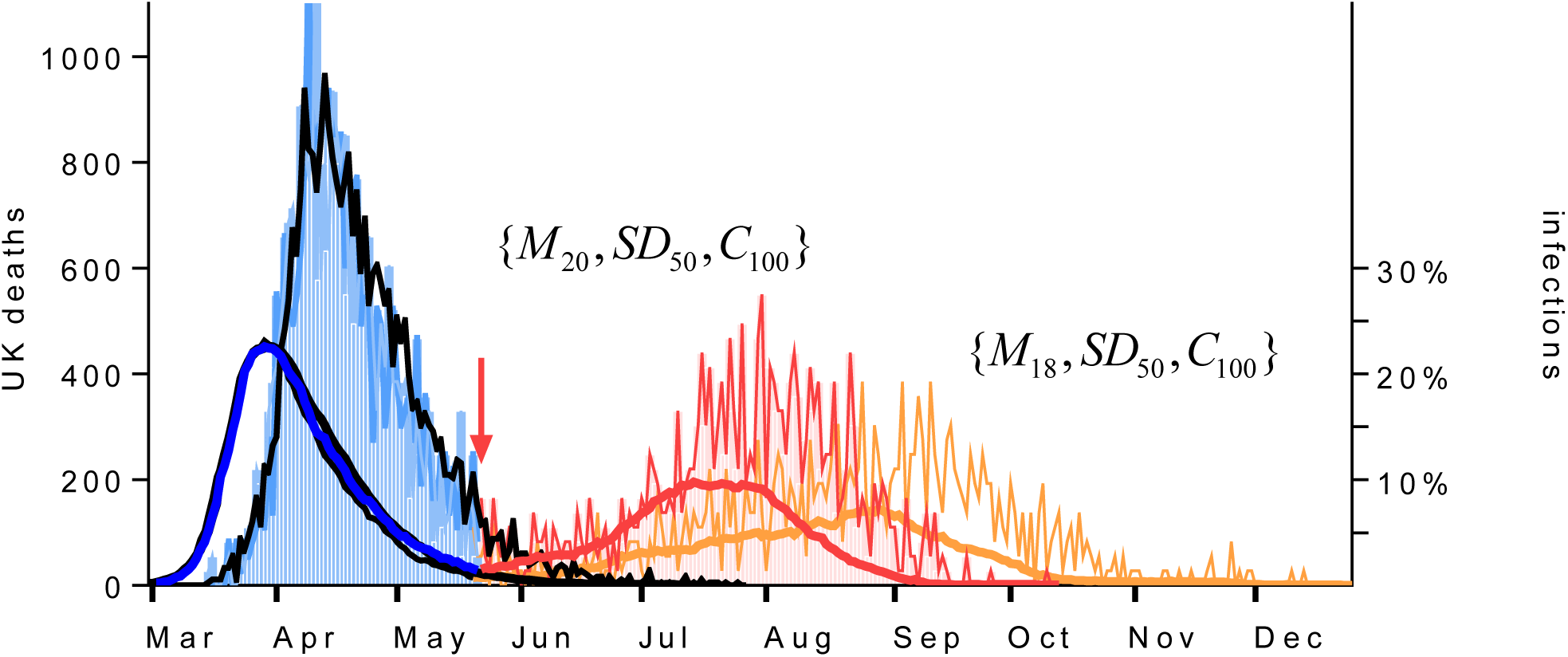
Relaxation of the lockdown on the 21^st^ May allows only an increase in movements from 7 to 18 in orange and from 7 to 20 in red whilst maintaining social distancing and confining the symptomatic.

The final illustration is of a relaxation of restrictions on the 1^st^ July, going to 20 movements a day, with social distancing being achieved in only 30% of interactions under 2 metres, and confining only 75% of the symptomatic. These parameters are sufficient to generate a second peak in the half of the population who are still susceptible, see Figure 12.

**Figure 12.**
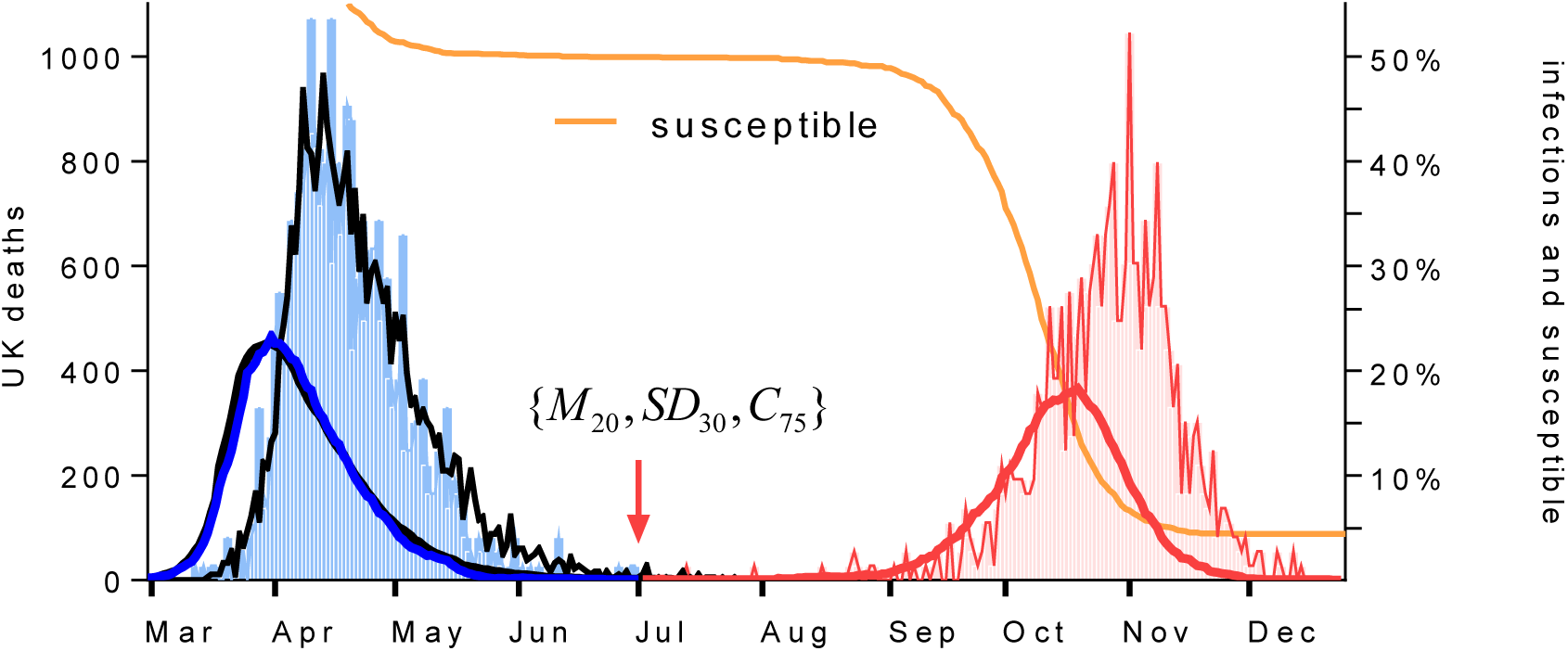
In black the locked down epidemic. In blue a lockdown epidemic with relaxation of the lockdown on the 1^st^ July after which the epidemic is shown in red. This relaxion represents a total of 57,000 UK deaths.

There are two epidemics in Figure 12 linked by only one infected individual. Repeated simulations did not always generate a second peak. This is a perfect example of a chaotic bifurcation. In other simulations, maintaining *M*_18_ and *C*_75_ but with *SD*_40_*, SD*_50_*, SD*_60_ there was no second-wave. Social distancing was sufficient to prevent a second epidemic in this situation.

In the final illustration, the pre-mitigation parameters {*M*_18_*, SD*_10_*, C*_90_} were used to generate an epidemic without mitigations, in search of the 260,000 deaths modelled by Imperial College (1). The epidemic was impressive, see Figure 13.

**Figure 13.**
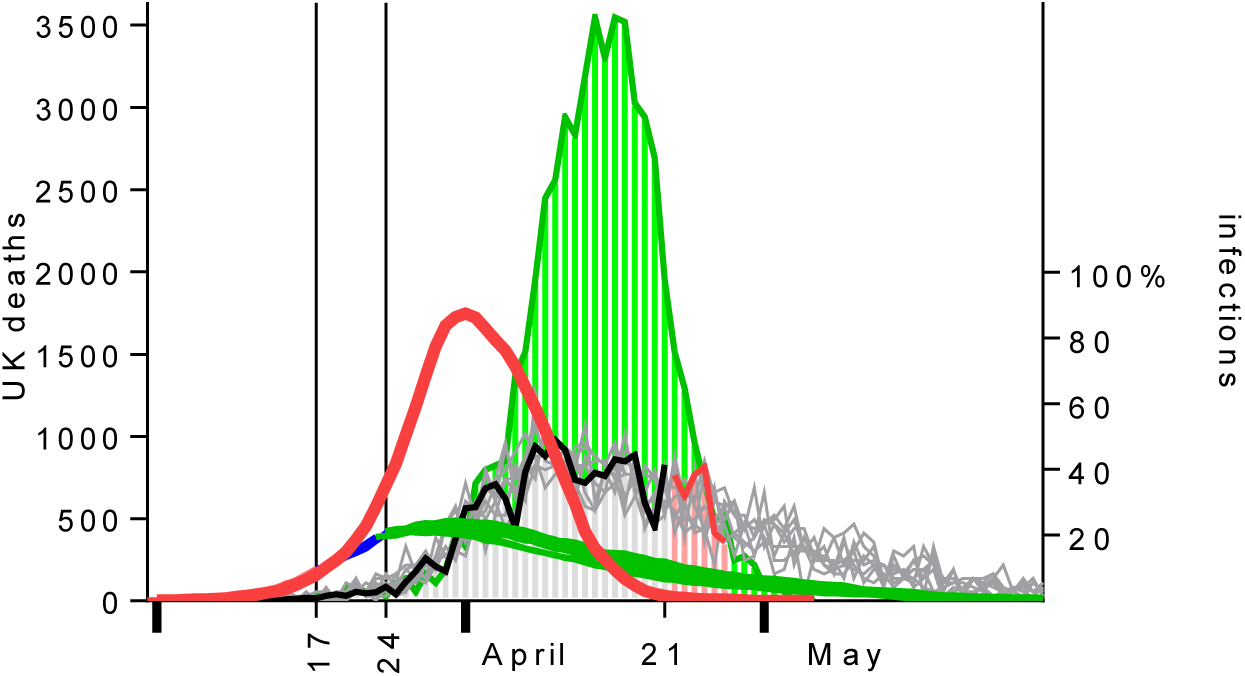
In red, an epidemic without any mitigations (right axis), initially doubling every three days and without mitigating interventions. In green, the associated and subsequent mortality. The rest of the Figure is taken from Figure 5.

The two month-long epidemic shown in Figure 13, which experienced no mitigating interventions, would have accounted for around 65,000 deaths in this analysis, somewhat short of the 260,000, but nonetheless a challenge in its magnitude and brevity. The unmitigated epidemic is shown in contrast to the achievements of social distancing and the lockdown which ‘flattened the curve’.

## Discussion

It is well-recognised that measuring mortality is surprisingly complex, but the challenges therein should not deter us from examining the shape of the UK Covid-19 mortality data. It should be examined on its own and in the context of the two mitigating interventions, which will have left their impressions on that shape. There is a lot of information encoded within the mortality data. This work set out to recover that information.

This author initially set out expecting to find, and hoping to confirm that, stochastic processes and chaotic dynamics would be sufficient to challenge the concept of a second-wave. The author then believed that the initial rapid exponential phase of unmeasured infection would have had to have been so great to create a 3-day doubling in deaths 20 days later, that the remnant susceptible population would be insufficient to maintain a second epidemic. This work did not set out to fit curves to support a hypothesis for the existence of a second-wave, but rather it set out to disprove their possible existence for the UK Covid-19 epidemic. Having now modelled many epidemics the author has been surprised at the findings and impressed with the impact of the lockdown.

The lockdown is crippling the economy and has other deleterious impacts on health. It is tempting to lift it now deaths are falling but this work supports prevailing arguments that a staged and gentle relaxion of the restrictions should take place. The simulations presented here demonstrate that maintaining social distancing during a relaxation of restrictions would be the most effective mechanism for limiting spread and containing a second-wave. However, this conclusion is almost inevitable since social distance through the inverse square relationship is central to this model’s mechanism of transmission.

The absolute number and nature of the movements reported in this work should not be overinterpreted as having any implication about the actual number and nature of real-life social interactions and how they might be modified. This model simulates *more* or *fewer* interactions and *closer* or *more separated* interactions and nothing more than these. The number of movements, their magnitude and their variance were introduced into the model purely as a device to permit experimentation to establish adjustable parameters capable of finding epidemics with the infected population growing in a similar way to that suggested by Covid-19 data and which permitted second-wave phenomena for study.

Some consider that asymptomatic transmission might not be a major driver of transmission (2). Asymptomatic infection has been considered to be relatively rare but many cases who are asymptomatic on the date of identification have gone on to develop disease and so the proportion of truly asymptomatic infections remains unclear. There is some asymptomatic transmission in this model which could be increased.

Individuals who remained susceptible after the first peak in infections had slightly higher susceptibility factors making them slightly less susceptible (data not shown). The second peak represented slightly less vulnerable individuals in these simulations. In the real-world something similar may also apply.

In this model neither the number of movements nor the distances moved were stratified according to age or sex. It is tempting to incorporate them to simulate, for example, a greater number of interactions between children. Such refinements might model the impact of allowing children to return to school as part of a staged relaxation of social restrictions. However, it is likely that the noise generated by the stochastic mechanisms would make such nuances difficult to detect. There is always a risk in over-elaboration or over parameterizing an otherwise simple and robust model.

## Data Availability

All data is referenced

## References

1. Mahase E. Covid-19: UK starts social distancing after new model points to 260 000 potential deaths. BMJ [Internet]. 2020 Mar 17 [cited 2020 Apr 22];m1089. Available from: http://www.bmj.com/lookup/doi/10.1136/bmj.m1089

2. WHO. Report of the WHO-China Joint Mission on Coronavirus Disease 2019 (COVID-19) [Internet]. 2020. Available from: https://www.who.int/docs/default-source/coronaviruse/who-china-joint-mission-on-covid-19-final-report.pdf

3. Wu JT, Leung K, Leung GM. Nowcasting and forecasting the potential domestic and international spread of the 2019-nCoV outbreak originating in Wuhan, China: a modelling study. The Lancet [Internet]. 2020 Feb 29 [cited 2020 Apr 5];395(10225):689–97. Available from: https://www.thelancet.com/journals/lancet/article/PIIS0140-6736(20)30260-9/abstract

4. Box, G.E.P. Robustness in the strategy of scientific model building. In: Launer, R. L, Wilkinson, G. N, editors. Robustness in Statistics. Academic Press; 1979. p. 201–36.

5. Lauer SA, Grantz KH, Bi Q, Jones FK, Zheng Q, Meredith H, et al. The incubation period of 2019-nCoV from publicly reported confirmed cases: estimation and application. medRxiv [Internet]. 2020 Jan 1;2020.02.02.20020016. Available from: http://medrxiv.org/content/early/2020/02/04/2020.02.02.20020016.abstract

